# Real World Performance of SARS-CoV-2 Antigen Rapid Diagnostic Tests in Various Clinical Settings

**DOI:** 10.1101/2021.03.02.21252400

**Authors:** Gili Regev-Yochay, Or Kriger, Sharon Beni, Carmit Rubin, Michael J. Mina, Bella Mechnik, Sabrina Hason, Elad Biber, Bian Nadaf, Yitshak Kreiss, Sharon Amit

## Abstract

**Background:** Several uses of Antigen rapid diagnostic tests (Ag-RDT) have been suggested. Analytical studies reported high specificity yet with lower sensitivity for detecting SARS-CoV-2 compared to qRT-PCR. Here, we present the use of these tests as a decision support tool in several settings.

**Methods:** Samples were collected for both Ag-RDT and qRT-PCR in three different clinical settings; 1. Symptomatic patients presenting at the Emergency Departments 2. Asymptomatic patients screened upon hospitalization and 3. Health-care workers (HCW) following SARS-CoV-2 exposure. Positive percent agreement (PPA), negative percent agreement (NPA), positive predictive value (PPV) and negative predictive value (NPV) were calculated. To estimate the association between Ct value, Ag-RDT and the number of days since SARS-CoV-2 exposure or symptomatic COVID-19, a mixed model was applied.

**Results:** A total of 5172 samples were obtained from 4595 individuals, with Ag-RDT and qRT-PCR results. Of these, 485 samples were positive by qRT-PCR. The PPA of Ag-RDT was greater for lower Ct values, reaching 93% in cases where Ct value was lower than 25 and 85% where Ct value was lower than 30. PPA was similar between symptomatic and asymptomatic individuals. The NPV and PPV were 96.8% and 99.1%, respectively. We observed a significant correlation between Ct value and time from infection onset (p<0.001). Lower Ct values were significantly associated with a positive Ag-RDT (p=0.01).

**Conclusions:** Ag-RDT can be used as a decision support tool in various clinical settings and play a major role in early detection of SARS-CoV-2 infected individuals, highly specific and with high sensitivity to the infectious stage of disease, whether symptomatic or asymptomatic.

## Introduction

Currently the benchmark standard test used for diagnosing and screening people suspected to be infected with SARS-CoV-2 is polymerase-chain-reaction (PCR). These tests have high analytic sensitivity, very high specificity but are very costly, time consuming and with relatively slow turn-around time (TAT)(1). To efficiently break infection chains, rapid results enabling fast isolation and contact tracing is required. This necessitates faster and less expensive diagnostic tests.

Multiple antigen rapid diagnostic tests (Ag-RDT) for SARS-CoV-2 have been recently developed(2–8) and few have received Emergency Use Authorization (EUA) from the US Food and Drug Administration (FDA). These tests are designed to directly detect SARS-CoV-2 proteins in respiratory secretions. These tests typically use a sandwich immunodetection employing lateral flow test format. The target is mostly the virus’ nucleocapsid protein. These tests are designed to be point-of-care (POC), rapid and inexpensive.

The World Health Organization (WHO)(6) has suggested several uses for Ag-RDT, including screening at-risk individuals in outbreak investigations, in order to rapidly isolate positive cases or screening front-line HCW, care homes, prisons & schools, to detect and isolate positive cases early. They also suggested these tests may be useful in clinical settings where PCR confirmatory testing is available. The European Center for Disease Prevention & Control (ECDC) support the use of Ag-RDT to increase COVID-19 testing capacity and state that these tests can help reduce further transmission through early detection of highly infectious cases, enabling rapid contact tracing (European Centre for Disease Prevention and Control. Options for the use of rapid antigen tests for COVID-19 in the EU/EEA and the UK. 19 November 2020. ECDC: Stockholm; 2020.).

While Ag-RDT have shown to have high specificity (>97%), they have been criticized to have sub-optimal analytic sensitivity compared to qRT-PCR detection of RNA. However, the need for a high analytic sensitivity test for the use of successfully containing the ongoing COVID-19 pandemic, has been questioned(9). More importantly, early and rapid detection of highly infectious individuals, i.e. asymptomatic, or presymptomatic patients as well as early symptomatic patients (during the first days of symptoms), when viral load is typically high, should be the prior target. Thus, sensitivity for infectious individuals, rather than analytic sensitivity against detection of potentially non-infectious RNA is important for this aim. Because the “ground-truth” is set as the result of a gold-standard which itself is imperfect, we discuss accuracy of the Ag-RDT not in terms of overall biological sensitivity and specificity, but instead in terms of percent agreement: negative percent agreement (NPA) and positive percent agreement (PPA) indicate the fraction of negative and positive RT-PCR results, respectively, that are repeated on the Ag-RDT.

Here, we present our initial real-world experience with several rapid antigen detection kits in a large tertiary medical center in Israel, utilizing the kits for Emergency Department (ED) triage, as well as in early detection of asymptomatic cases, mainly in support of outbreak investigations among HCW, ensuring operational continuity despite high prevalence of COVID-19 infections in the community.

## Methods

### Setting

The Sheba Medical Center is the largest tertiary hospital in Israel, with 1600 beds, of these, 1400 acute care beds. Over 9000 HCW are occupied, of these, 1770 physicians and 3124 nurses.

### Study population and Ag-RDT testing strategies

Three major testing strategies were used in this study: 1) As a decision support tool in the patient triage process, for a cohort of symptomatic patients, mostly visiting ED, who were suspected as being infected with SARS-CoV-2 **(Figure 1)** 2) As a strategy for early detection of asymptomatic cases among two cohorts: i) asymptomatic patients screened for COVID-19, including women before delivery arriving to the obstetric ED (OB-ED), and ii) a cohort of HCW, who were screened following exposure to a detected COVID-19 case. In addition, for a small subset, these tests together were used as a decision support tool, to allow COVID-19 recovered HCW to return to work even if qRT-PCR was still positive (For mild infections with at least 10 days post-infection and asymptomatic for at least 3 days).

**Fig 1.**
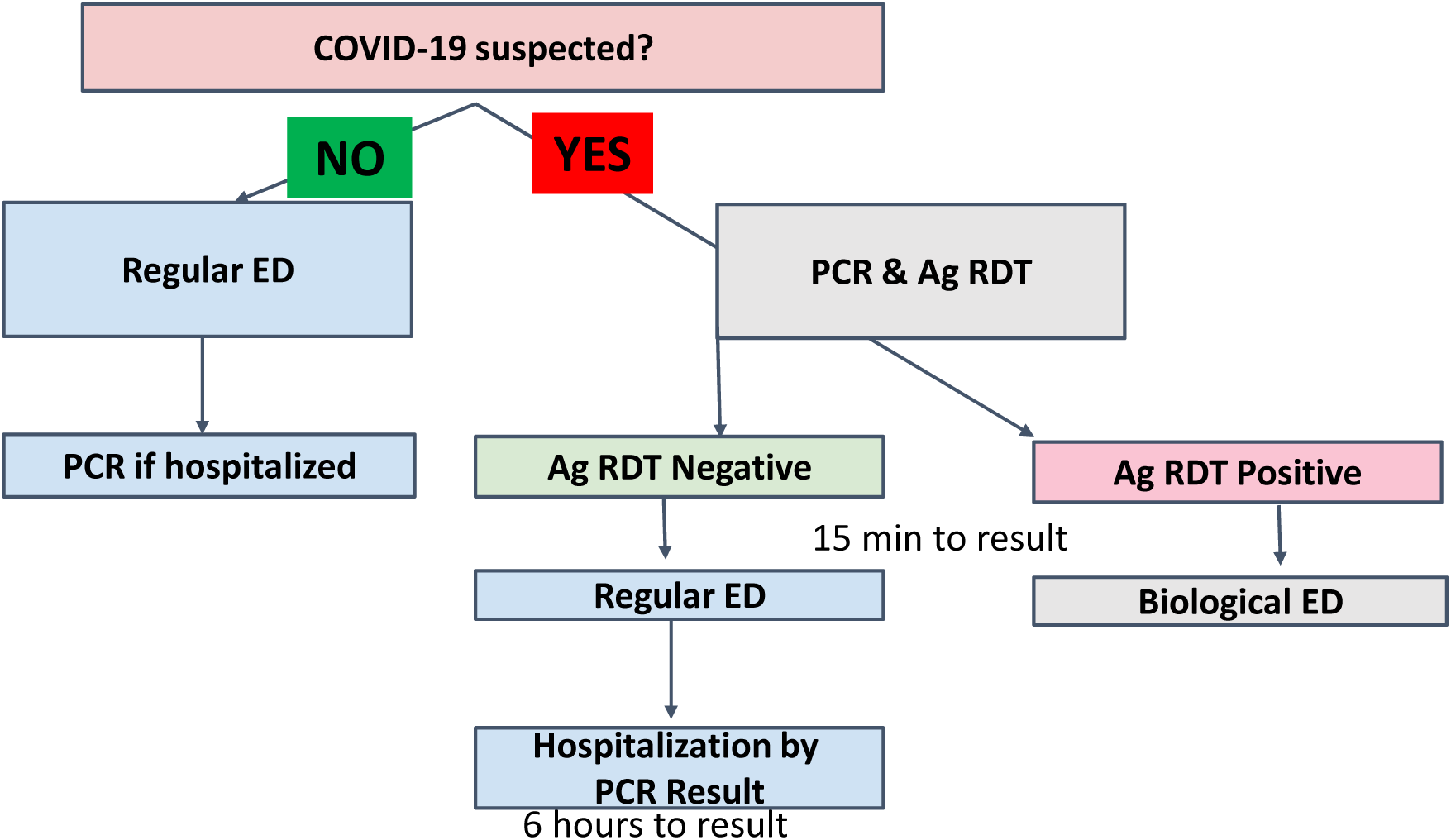
Testing strategy in the Emergency Department.

### Sample collection

a nasopharyngeal nasal swab sample was collected by trained personnel following appropriate safety precautions and used in various Ag-RDT kits (Nowcheck COVID-19 Ag test (Bionote, S. Korea)(10), Panbio™ COVID-19 Ag rapid test, (Abbott Rapid Diagnostic Jena GmbH, Jena, Germany)(2), BD Veritor™ (Becton, Dickinson and Company Franklin Lakes, NJ)(8), GenBody COVID-19 Ag (GenBody Inc, S. Korea)(11), STANDARD Q COVID-19 (SD-Biosensor Inc, S. Korea)(11), following the manufacturer’s instructions (**Supplementary Table**). In addition, naso-and oropharyngeal samples were obtained and tested for SARS-CoV-2 quantitative real-time polymerase chain reaction (qRT-PCR) test. For qRT-PCR, NP swabs were placed in 3mL of universal transport medium (UTM) or viral transport medium (UTM) and test was performed according to manufacturers’ instructions on various platforms: Allplex™ 2019-nCoV (Seegene, S. Korea), NeuMoDx™ SARS-CoV-2 assay (NeuMoDx™ Molecular, Ann Arbor, Michigan), Xpert®, Xpress SARS-CoV-2 (Cepheid, Sunnyvale, CA, USA).

### Data collection

Patient information was gathered through the hospital electronic database. Data included socio-demographic details, as well as duration, type of symptoms and laboratory tests including previous or proceeding SARS-CoV-2 qRT-PCR tests and SARS-CoV-2 IgG antibody levels when available. Day of COVID-19 infection onset was determined either by the first positive SARS-CoV-2 qRT-PCR or first day of symptoms, whichever was earlier. qRT-PCR cycle threshold (Ct) values of the N-gene were used as a correlate of SARS-CoV-2 viral load.

### Statistical analysis

PPA, NPA, PPV and NPV were calculated for Ag-RDT compared with qRT-PCR results.

To estimate the association between Ct value, Ag-RDT and the number of days since SARS-CoV-2 infection (defined as the onset of symptoms or first detection), a mixed model was applied with the log transformed Ct value as the outcome. A random intercept was allowed for each subject to account for the correlation between repeated measurements of the same subject, assuming a similar correlation between each pair of measurements, i.e. Compound Symmetry. The model included number of days since SARS-CoV-2 infection as well as log transformed days, to capture the non-linear association between Ct value and time since infection, either as defined by symptoms or by first positive qRT-PCR. The predicted association as well as its 95% confidence interval (CI) were calculated using the exact binomial method; p-values <0.05 were considered statistically significant.

A generalized linear mixed regression model was used to investigate independent predictors for a positive Ag-RDT test. Variables considered in the model included: whether the patient was symptomatic or asymptomatic, the number of days from disease onset to positive Ag-RDT(less or greater than 10 days), and Ct value (divided into 3 categories: lower then 30, 30-35 and greater than 35). Similar to the mixed model described above, we allowed a random intercept for each subject.

### Ethical committee approval

The research protocol was approved by the Sheba Institutional Review Board and oral informed consent was obtained from study participants.

## Results

A total of 5172 samples were obtained from 4595 individuals who were tested by both Ag-RDT and qRT-PCR, including 3594 samples from symptomatic patients arriving to the ED, patients hospitalized in COVID-19 wards or symptomatic HCW, and 1548 samples from asymptomatic patients, with no previous COVID-19 detection. In addition, 30 samples were collected from 26 recovered COVID-19 patients of whom 25 had a positive qRT-PCR and one had a positive Ag-RDT (**Figure 2**). Further demographic data is shown in **Table 1**.

**Table 1:** Study population

|  |  | Total |  | Symptomatic |  | Asymptomatic |  |
| --- | --- | --- | --- | --- | --- | --- | --- |
|  |  | N | % | N | % | N | % |
| Total number of individuals tested |  | 4827 |  | 3429 |  | 1398 |  |
| Total number of samples |  | 5142 |  | 3594 |  | 1548 |  |
| Sex (Male) |  | 2026 | 41.97% | 1751 | 48.72% | 275 | 17.76 |
| Age | Mean | 56.7 |  | 61.9 |  | 43.8 |  |
|  | Median | 50.1 |  | 61.0 |  | 36.4 |  |

**Figure 2:**
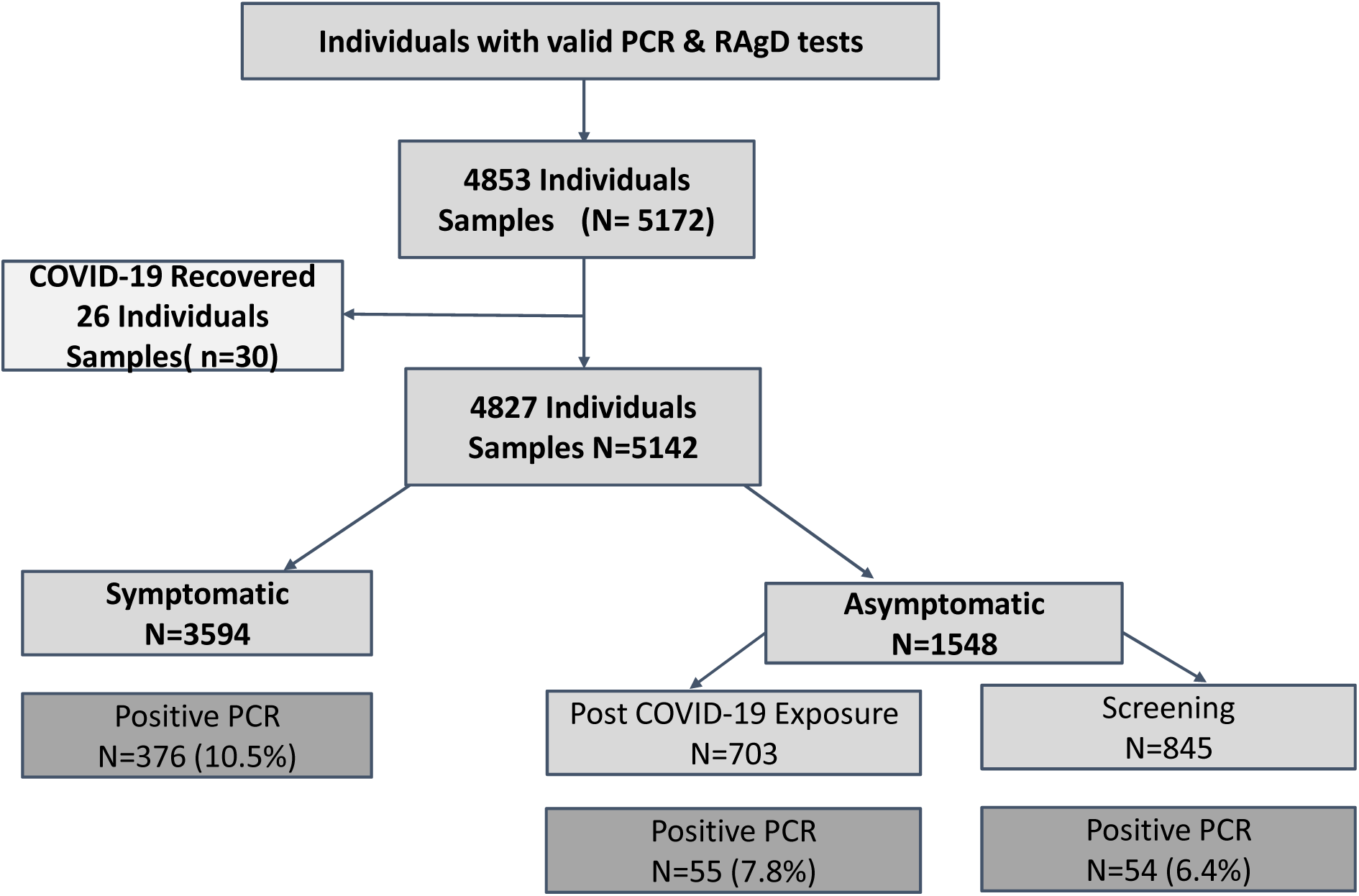
Study population.

Of all samples collected, 4659 were negative for SARS-CoV-2 by qRT-PCR. Three of 4659 negative qRT-PCR samples had a positive Ag-RDT, thus the overall NPA was 99.92%. A positive qRT-PCR was detected in 485 cases. In 79 (15%) cases, the samples were collected from patients who had a positive qRT-PCR or COVID-19 symptom onset starting greater than the preceding 10 days.

The median Ct value of all 485 positive samples was 26 (interquartile range (IQR): 20.04-32.18). The median Ct was significantly higher for cases where Ag-RDT was negative (median 32.58, IQR: 28.9-35.75), vs. cases where it was positive (median 21.97, IQR: 18.1-26.8).

The PPA between Ag-RDT and qRT-PCR was greater for lower Ct values, reaching 93% in cases where Ct value was lower than 25. The overall PPA for all cases where Ct value was 35 or less, was 74% and negative predictive value (NPV) was 96.8% and positive predictive value was 99.1%. When only newly detected cases were included, i.e. diagnosed within the preceding 10 days, overall sensitivity was 76% (for Ct<35), 85% for those with Ct<30 and increasing to 93% for those with Ct<25. (**Table 2a**).

**Table 2:**
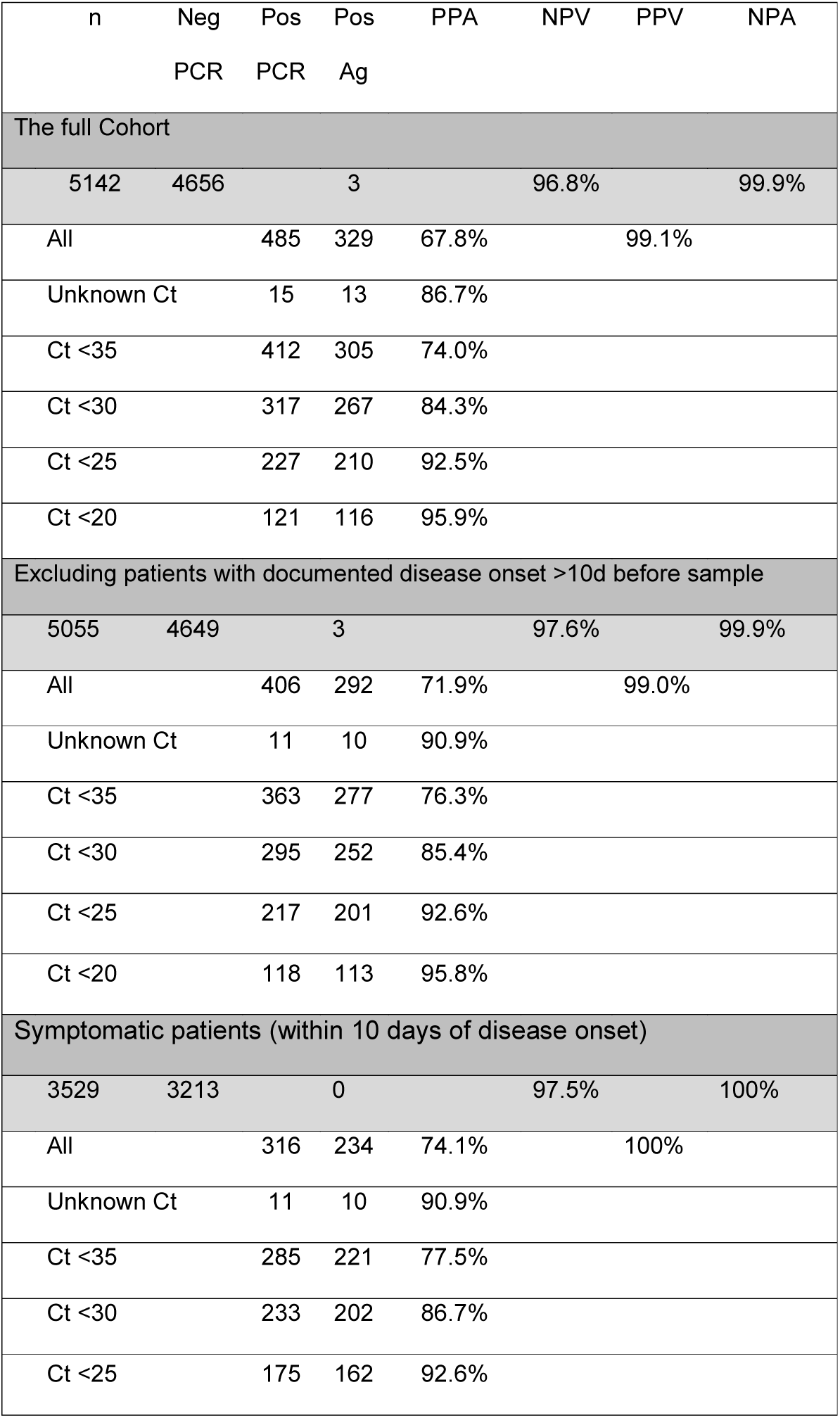
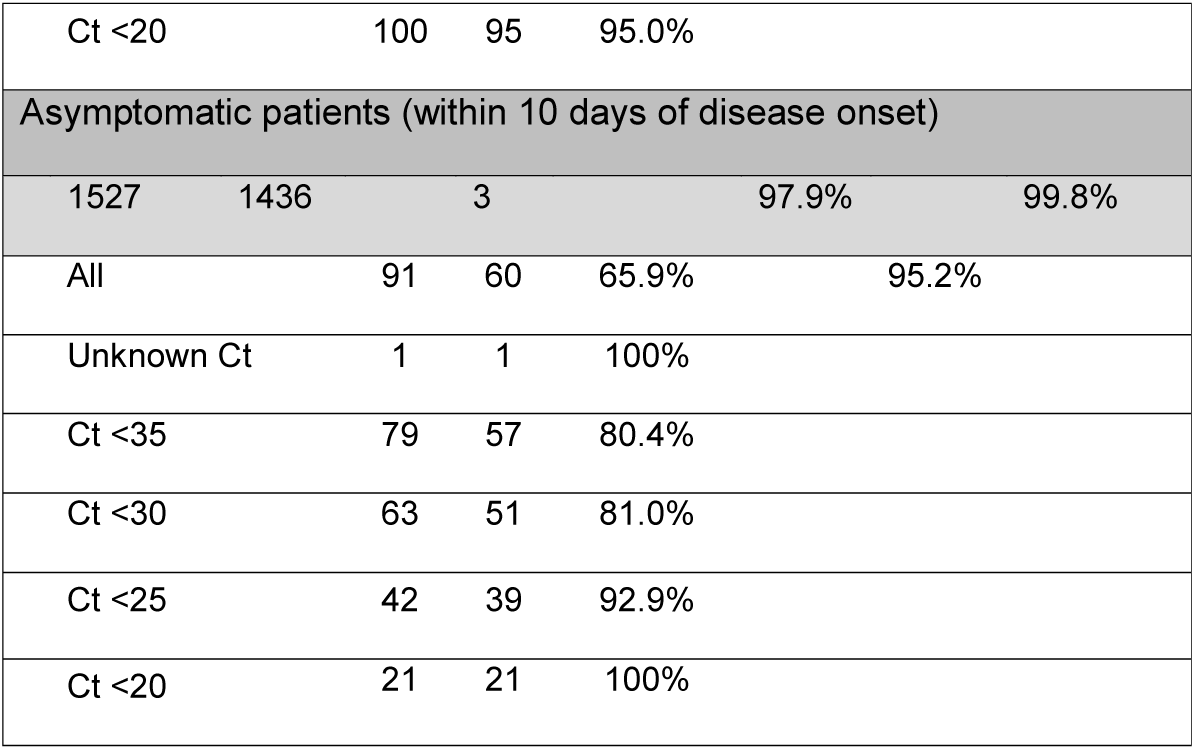
PPA, NPA, NPV and PPV

While symptomatic patients were older, mainly patients arriving at the ER, and asymptomatic patients were mainly younger HCW, or screened obstetric patients, the median Ct amongst the positive detected samples was not significantly different (24.9, IQR 19.2-31.4 vs. 26.7, IQR 21.5-33.3, respectively). The PPA was also similar amongst the two groups (**Table 2b**).

In agreement with the above, a generalized linear mixed regression model identified that a positive Ag-RDT was significantly associated with lower Ct values (OR=0.04, 95%CI 0.018-0.095, P<0.0001 for Ct value greater than 35) and older age, but not with symptomatic presentation (**Table 3**).

**Table 3:** Predictors of positive Ag-RDT multiple logistic regression:

|  |  | Adjusted<br>OR | 95% CI | P-value |
| --- | --- | --- | --- | --- |
| Time from<br>infection | 0-10d |  |  |  |
|  | 11+ d | 0.356 | 0.09-1.28 | 0.098 |
| Ct value |  |  |  | 0.012 |
| Symptomatic | Vs.<br>Asymptomatic | 0.383 | 0.995-1.06 | 0.285 |
| Gender | Male vs.<br>Female | 1.263 | 0.384-4.16 | 0.657 |

When we characterized the 87 false negative cases diagnosed within the 10 days of disease onset in which Ct value was lower than 35, we found that 7 of these had been diagnosed for over a week, but 73 cases were newly identified cases (days 0-3 following diagnosis either by symptom onset or first positive qRT-PCR test). Of these 73 cases, 60 were identified on the test day (day 0). Owing to limited pre-detection data, it was unclear if these individuals are new infections or individuals with detectable RNA but no infectious virus remaining in their nasal passage. To determine if most missed cases were early versus late infection, we assumed that if, on repeated testing, viral RNA load was dropping (i.e. higher Ct values) then virus was likely in the clearance phase at time of first detection. For 18 of the 60 cases, we had follow up qRT-PCR tests with Ct values and could estimate which were recovering and which were truly new cases. Recovery was defined as consecutive tests on day1-3 with stable or increasing Ct value, and new cases as those with consecutive decreasing Ct values. Of the 18 cases for which we had day 0 and follow up data, 12 (67%) were recovering patients (**Figure 3**).

We observed a significant correlation between Ct value and time from infection onset; A mixed model using repeated measures with a random intercept for each subject indicated a significant non-linear relation between Ct value, and the log of number of day post SARS-CoV-2 infection (p<0.0001). The variance of the observed Ct values was greatest on the day of SARS-CoV-2 diagnosis. During the first 10 days following the diagnosis, a logarithmic increase in Ct value was observed followed by a plateau. Following 20 days from infection, all cases had low viral loads, as defined by Ct value>30, apart from a single case of a patient with severe immunodeficiency (Common Variable Immuno-deficiency CVID) (**Figure 3**). A positive Ag-RDT was observed only during the first 20 days and in 89% within the first ten, with a single exclusion of the CVID case, in which the test was also positive.

## Discussion

We present real world use of SARS-CoV-2 Ag-RDT as an immediate clinical decision support tool in several clinical settings. RDT’s had previously revolutionized the diagnosis and treatment of infectious diseases, most notable are malaria and streptococcal throat infections(12,13). In both these cases rapid diagnostics, performed by the primary care taker as a triage measure, may allow for a prompt rule-in/rule-out decision. Despite a relatively low analytic sensitivity compared to PCR, the advantages of RDT’s are imminent: rapid result, simple operation and handling (size of kits, storage, use) allowing for point-of-care performance and minimal training, and low pricing. Large international health authorities (WHO, CDC, ECDC) have published papers referring to the use of RDT’s for COVID-19 as a testing/screening tool in various settings, however real-life data regarding it performance are lacking(5).

In this study RDT’s of various manufacturers were used for two main purposes – at the ED, as a triage measure for improving patient-flow and placement, and as a strategy for early detection of asymptomatic cases, at the personnel clinic for HCW following suspected exposures and at the obstetric ED for screening women before labor. In all settings, we tested patients/HCW with no symptoms or symptoms of various lengths, a design enabling an unbiased analysis of the performance of RDT’s.

As expected, specificity of all kits is very high (>99.7%), reassuring the strong positive predictive value of the assays, especially during peaking incidence. This may allow for prompt decision in all our study settings. At the ED – as an additional triage tool, allowing early differentiation between SARS-CoV-2 infectious patients and other suspected patients, allowing discharge of mild cases to home-care and isolation, or hospitalization in COVID-19 wards in more severe cases. At the obstetric ED, as a screening tool, allowing early detection and isolation of asymptomatic COVID-19 infectious patients. And last, at the personnel clinic – early detection of positive HCW, allowing exposed HCW to continue to work and avoid unnecessary isolation when Ag-RDT is negative and rapid and early epidemiological investigation, without awaiting PCR result, when Ag-RDT is positive.

As for the negative predictive value, in concordance with previous publications(2–4), the overall analytical sensitivity of the kits is inferior to PCR, and correlates with the presumed viral load, represented by the Ct value (Table 2.). However, the correlation between kit performance, Ct and the time since the onset of symptoms resembles the current knowledge regarding infectivity kinetics with respect to viral load and time(14–16). Moreover, the direct correlation between positive Ag-RDT and infectivity have been recently reported (8). Thus, for a patient with a given Ct value, there is a lower chance of being infective the more time elapsed since symptoms onset. This also explains the pattern of sensitivity gaps between symptomatic and asymptomatic cases, with similar sensitivity with high viral loads (Ct<25) yet lower in asymptomatic patients with lower viral loads (Ct>30). As most published data show similar infectiveness regardless of symptoms, and Ag-RDT’s detect viral particles in the nasopharyngeal cavities, symptoms merely create a lead-time bias, allowing for early detection. Ag-RDTs perform as well in both symptomatic and asymptomatic patients, as has been recently described (17).

Taken together, as the viral-load curve of SARS-CoV-2 shows a short, sharp incline and a long, moderate decline, with a relatively short infectious period with respect to PCR positivity(9,18) our findings suggest that at a given time point, an RDT-negative PCR-positive patient, will likely present a non-infective individual.

The study had several limitations. First, day of infection onset was defined as either day of first positive qRT-PCR, or day of symptoms onset, yet for some patients this may not be the precise day of infection, and we suspect that many of those diagnosed on day 0, were actually already infected for several days (19). This may particularly be frequent for the asymptomatic patients, whom are likely more frequently recovering and no longer infectious by the time they are detected(17). Second, we did not test a single specific Ag-RDT, but rather combined the results of several kits. This was done, since our objective was to show the potential use of the method, rather than the performance of a specific kit. But since the performance of each kit in similar settings did not differ significantly, we believe that for our objective this is not a significant limitation.

In conclusion, this report of a real-life experience with SARS-CoV-2 RDT’s offers several settings in which both its PPV and NPV prove of value. The very high specificity enables immediate rule-in of positive SARS-CoV-2 infectious cases, be it symptomatic ED patients, exposed or early-recovered HCW. Despite the lower analytic sensitivity, accumulating bulk of data support Ag-RDT as a better surrogate for infectivity than PCR, as it represents translated viral proteins rather than RNA remnants. For example, Pekosz et al. (8) shows a better correlation between infectivity and Ag-RDT+/PCR+ cases than Ag-RDT-/PCR+ cases. In a hospital setting, an Ag-RDT is always backed by a PCR which will minimize the chance of missing a very early disease in the case of presymptomatic or asymptomatic patients. However, our data support its prudent use as a rapid decision-support tool allowing for an efficient ED flow and management of hospital staff during peaking COVID-19 prevalence.

## Data Availability

All data is available at Sheba Medical Center

## Acknowledgements

We would like to thank Efrat Steinberger for coordinating the study, Amir Grinberg and Amit Gotkind for administrative and management assistance.

---

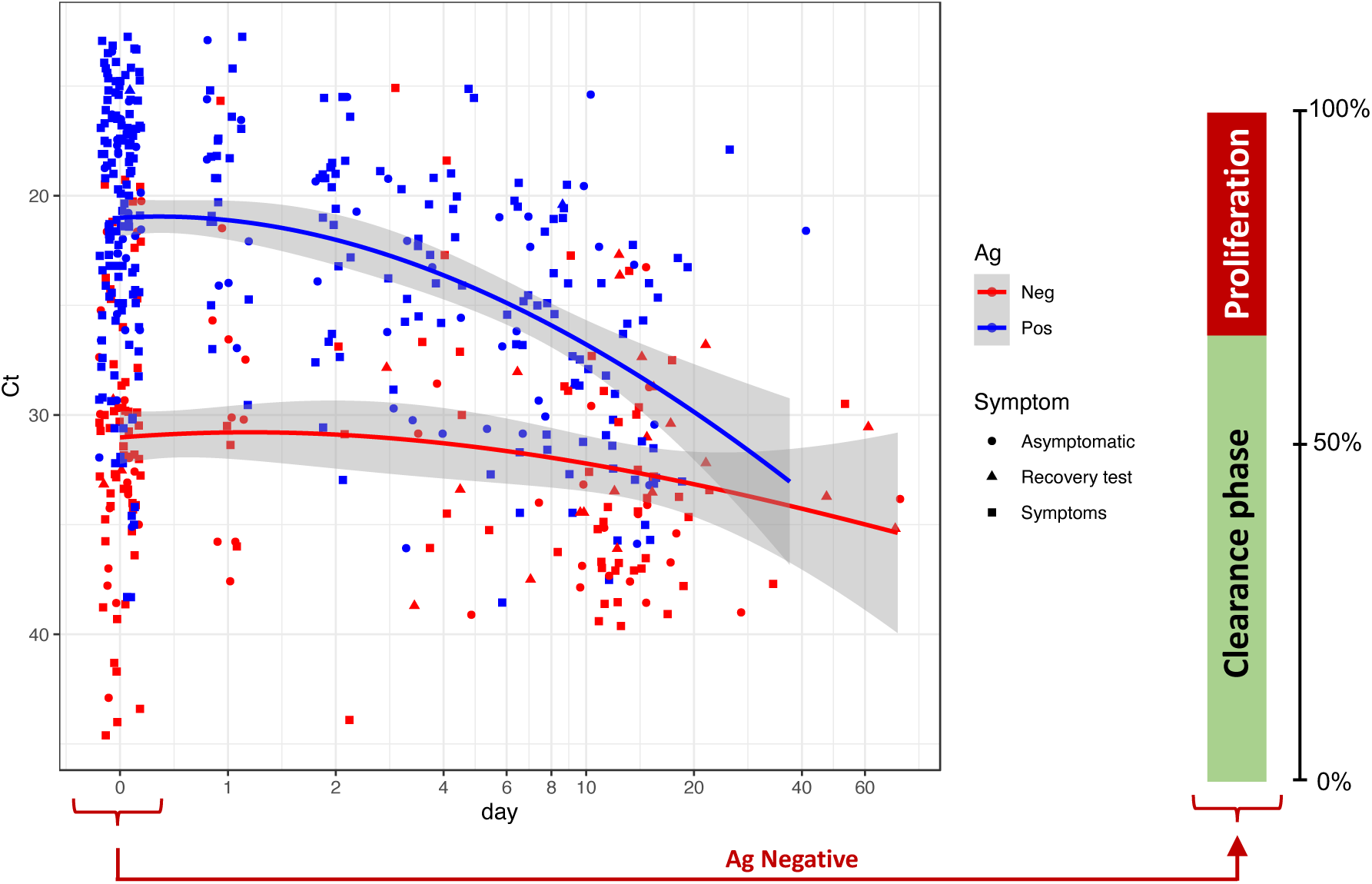

