## Supplementary material for "Real World Performance of SARS-CoV-2 Antigen Rapid Diagnostic Tests in Various Clinical Settings": Supp Table

Supplementary Table

| **RAD test name** | **Manufacturer** | **Number of samples** |
| --- | --- | --- |
| Nowcheck COVID-19 Ag test | Bionote, S. Korea | 3038 |
| Panbio™ COVID-19 Ag rapid test | Abbot laboratories, Germany | 582 |
| BD Veritor™ | Becton, Dickinson and Company Franklin Lakes, NJ | 150 |
| GenBody COVID-19 Ag | GenBody Inc, S. Korea | 207 |
| STANDARD Q COVID-19 | SD-Biosensor Inc, S. Korea | 288 |
